# Beta activity reflects change in upper limb activity rather than impairment following high-dose high-intensity upper limb neurorehabilitation in chronic stroke

**DOI:** 10.64898/2026.03.19.26348794

**Authors:** Catharina Zich, Sebastian Sporn, Lisa Tedesco Triccas, Mireia Coll I Omana, Sven Bestmann, Nick S Ward

**Affiliations:** Department of Clinical and Movement Neuroscience, UCL Queen Square Institute of Neurology, United Kingdom; Wellcome Centre for Integrative Neuroimaging, FMRIB, Nuffield Department of Clinical Neurosciences, University of Oxford, United Kingdom; Medical Research Council Brain Network Dynamics Unit, University of Oxford, United Kingdom; Faculty of Rehabilitation Sciences, REVAL Rehabilitation Research Center, Hasselt University, Diepenbeek, Belgium; Center for Clinical Neuroscience, Hospital Universitario Los Madroños, Brunete, Madrid, Spain; Wellcome Centre for Human Neuroimaging, Department of Imaging Neuroscience, UCL Queen Square Institute of Neurology, United Kingdom; The National Hospital for Neurology and Neurosurgery, Queen Square, London WC1N 3BG, United Kingdom

**Keywords:** Arm, Chronic stroke, mobile EEG, Recovery, Rehabilitation

## Abstract

**Background:** High-dose high-intensity upper limb neurorehabilitation can lead to meaningful clinical gains even in chronic stroke, yet substantial variability in recovery remains unexplained. Identifying neurophysiological markers linked to neuroplasticity and recovery could provide mechanistic insights and guide personalised rehabilitation.

**Objective:** To characterise stroke-related alterations in β-activity during movement and neural activity at rest and explore associations between brain activity and changes in upper limb clinical outcomes in chronic stroke survivors undergoing three-week high-dose rehabilitation.

**Methods:** Electroencephalography (EEG) was recorded during the three-week rehabilitation programme in 40 chronic stroke survivors participating in the Queen Square Upper Limb (QSUL) Programme, as well as in 26 healthy controls. Recordings were taken during passive movement of the affected and unaffected index fingers (∼70 movements per hand) and at rest (∼7 min). Clinical assessments included the Fugl-Meyer Upper Limb Assessment (FM-UE), reflecting impairment-level deficits, and the Chedoke Arm and Hand Activity Inventory (CAHAI-13), capturing real-world upper limb activity, to examine their differential relationships with movement-related β-activity.

**Results:** Stroke survivors showed significant improvements in FM-UE and CAHAI scores following the rehabilitation programme (Mean Δ: FM-UE = 7.5, CAHAI = 7.4), exceeding minimum clinically important differences. Compared to controls, stroke survivors exhibited less strong β-event-related desynchronization/synchronization (β-ERD/ERS) during passive movement of the affected and unaffected index finger, with effects lateralised to the lesioned hemisphere. No significant differences at rest were observed between stroke participants and healthy controls. Only improvements in CAHAI, but not FM-UE, were associated with stronger β-ERD (more negative) and stronger β-ERS (more positive) responses during passive movement.

**Conclusions:** Stronger movement-related β-activity is associated with improvements in upper limb activity following high-dose high-intensity neurorehabilitation, suggesting β-activity as a potential marker of neuroplasticity.

## 1. Introduction

Stroke remains a leading cause of long-term adult disability worldwide, with persistent upper limb (UL) dysfunction and impairment among the most common and disabling consequences (Feigin et al., 2023). While most spontaneous motor recovery occurs within the first three months post-stroke (Hayward et al., 2021; Lee et al., 2015), around 50% of patients in the chronic phase (>6 months) continue to experience UL dysfunction and impairment (O’Flaherty & Ali, 2024), affecting independence and quality of life (Broeks et al., 1999). Evidence suggests that high-dose high-intensity neurorehabilitation can still produce meaningful improvements, even years post-stroke (Ward, 2017; Ward et al., 2019). However, there remains substantial inter-individual variability in outcomes (Johnstone et al., 2022; Ward et al., 2019). While baseline clinical status and rehabilitation dose are the strongest predictors of treatment response, they do not fully explain the variability in recovery trajectories (Salvalaggio et al., 2023).

Understanding the neural mechanisms underlying recovery in chronic stroke could inform targeted rehabilitation strategies. Electroencephalography (EEG) offers a non-invasive, high-temporal-resolution approach to study cortical activity linked to sensorimotor control. Movement-related sensorimotor β-activity (13–30 Hz) is reduced post-stroke (Laaksonen et al., 2012; Parkkonen et al., 2018; Tang et al., 2020; Zich, Mardell, et al., 2025; Zich, Ward, et al., 2025). Further, sensorimotor β-activity after stroke has been related to UL motor capacity (Laaksonen et al., 2012; Parkkonen et al., 2018; Tang et al., 2020), UL motor learning (Espenhahn et al., 2020; Rossiter et al., 2014) and UL motor recovery (Zich et al., 2017; Zich, Ward, et al., 2025). This aligns with the known roles of β-activity in motor inhibition, sensorimotor state maintenance, and interregional communication, as well as in cortico-muscular coherence reflecting functional coupling between motor cortex and muscle activity during movement (Engel & Fries, 2010; Kilavik et al., 2013; Mima & Hallett, 1999; Peng et al., 2024). Linking changes in β-activity to clinically meaningful improvements may provide mechanistic insight into recovery processes beyond what structural or baseline clinical measures alone can reveal.

To study such links, it is necessary to examine participants undergoing an intervention known to produce measurable clinical change. In this study, we recruited chronic stroke survivors participating in the Queen Square Upper Limb (QSUL) neurorehabilitation programme, a specialist NHS service delivering ∼90 hours of intensive UL training over three weeks (Kelly et al., 2020; Ward et al., 2019). The QSUL programme reliably induces clinically meaningful improvements in both UL impairment and activity (Ward et al., 2019), providing an ideal framework to investigate neural correlates of recovery in the chronic stage. Importantly, we measured recovery using two complementary clinical tools: the Fugl-Meyer Upper Limb Assessment (FM-UE), which primarily evaluates impairment-level deficits in motor control, and the Chedoke Arm and Hand Activity Inventory (CAHAI), which captures real-world UL activity (Barreca et al., 2005; Fugl-Meyer et al., 1975). This distinction allows us to test whether neural activity is differentially linked to activity outcomes versus impairment reduction.

In the present study, we recorded high-quality EEG at a single time point within the three-week rehabilitation programme during passive movement of the affected and unaffected UL, as well as at rest, alongside FM-UE and CAHAI assessments at admission and discharge. Our aims were to: i) assess stroke-related alterations in β-activity during passive movement of the affected and unaffected UL, with a focus on the limb-specificity of stroke-related effects; ii) assess stroke-related alterations in neural power at rest, examining the task-specificity of stroke-related effects; and iii) assess how β-activity during passive movement and neural power at rest relate to baseline clinical status and rehabilitation-induced change, across both impairment- and activity-based measures.

## 2. Materials and methods

### 2.1 Experimental design

#### 2.1.1 Ethical approval

The Research Ethics Committee of the University College London and the NHS Research Ethics Committee (London - Surrey Research Ethics Committee) approved the study protocol (20/LO/0520), and all subjects provided written informed consent.

#### 2.1.2 Subjects

Chronic stroke survivors ((≥ 6 months post-stroke) (Bernhardt et al., 2017)) were recruited from the QSUL programme between April 2022 and October 2023. QSUL is a UK NHS neurorehabilitation service delivering 90 hours of intensive UL rehabilitation over three weeks, designed to produce clinically meaningful improvements across a range of impairment and activity levels (Kelly et al., 2020; Tedesco Triccas et al., 2025; Ward et al., 2019). This well-characterised intervention provides an effective framework to examine relationships between behavioural recovery and neurophysiological measures (for TIDieR checklist see (Tedesco Triccas et al., 2025)). The inclusion criteria for this study were: aged 18 years or older; confirmed diagnosis of stroke with UL impairment; and no other brain injury, neurological condition or major psychiatric illness. Participants were excluded if they had a severe cognitive or communication impairment or were from a non-English speaking background when there was no translator available. Healthy controls (HC) were recruited via advertisements, reported no current psychiatric or neurological disorders, and were recruited to approximately match the age distribution of the stroke cohort.

#### 2.1.3 Clinical data

We focus on the modified upper-limb Fugl-Meyer Assessment (FM-UE) with a maximum score of 54 to assess stroke-specific performance-based impairment (Crow & Harmeling-van der Wel, 2008; Duncan et al., 1983; Fugl-Meyer et al., 1975; Hsieh et al., 2007). Page et al. (Page et al., 2012) reported an minimum clinically important differences (MCID) of 5.25 score for the standard 66-score FM-UE assessment in chronic stroke with minimal to moderate impairment. Additionally, we use the Chedoke Arm and Hand Activity Inventory (CAHAI-13), which has a maximum score of 91 and an MCID of 6.3 points (Barreca et al., 2005) to assess recovery of the UL whilst performing daily activities after stroke. These assessments capture different levels of the International Classification of Functioning, Disability and Health (ICF) framework. The FM-UE primarily measures impairment in the body functions and structures domain, whereas the CAHAI evaluates task performance in the activity domain. We use these assessments obtained on admission (T1) and discharge (T2), and calculated the normalised percentage 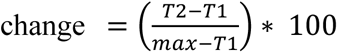. The normalised percentage change accounts for individual baseline differences and ceiling effects, allowing for a fairer comparison of improvement across patients by expressing change relative to each patient’s potential for recovery.

### 2.2 EEG data

#### 2.2.1 EEG data acquisition

Data were acquired under consistent conditions, with subjects seated comfortably in a chair in the same room within the hospital, for both chronic stroke patients and HC. EEG data were recorded with sintered Ag/AgCl electrodes from 30 scalp sites using electrode caps (Easycap, Herrsching, Germany), with AFz as the ground and FCz as the reference. Data were acquired with an amplitude resolution of 0.1 μV and a sampling rate of 500 Hz, using online analogue filter settings of 0.02 to 250 Hz, via a wireless amplifier (SmartingPRO, mBrainTrain, Belgrade, Serbia) attached to the back of the EEG cap. Data were transmitted via Bluetooth 5.0 to a laptop (Dell Latitude 7390) via a Bluetooth dongle (VCK dongle). No data were lost during transmission. Impedances were checked at the beginning (Smarting Streamer software v4.1; mBrainTrain, Belgrade, Serbia) and recorded throughout the experiment with a sampling rate of 125 Hz. Impedances were on average 16.9 kΩ (*SD* = 6.1 kΩ) in stroke survivors and 15.5 kΩ (*SD* = 5.9 kΩ) in HC.

#### 2.2.2 Passive movement task and resting state

Data were acquired during passive movements of the index fingers in a block design, considered affected or unaffected in stroke participants and dominant or non-dominant in HC. The order in which each UL was tested was counterbalanced across participants. We used a passive movement task as this provides access to movement-related β-activity (Parkkonen et al., 2015; Zich, Mardell, et al., 2025), independent of the individual’s movement capacity. The subject’s index finger was positioned above a keyboard key (Ecarke OSU Keypad 4-Key Gaming Keyboard) and held in place using a Velcro sleeve. The experimenter moved the key down and immediately up roughly every 5 s (Stroke survivors: *M* = 5.1 s, *SD* = 0.2 s; HC *M* = 5.1 s, *SD* = 0.1 s) for 6 min (Stroke survivors: *M* = 69.2, *SD* = 4.1, range 61-81 trials; HC *M* = 67.9, *SD* = 1.4, range 64-70 trials). Participants were instructed to relax, not to move, and not to pay attention to the passive movement.

Following the passive movement task, data were collected during rest with eyes open (Stroke survivors: *M* = 7.0 min, SD = 0.2 min; HC *M* = 7.0 min, *SD* = 0.1min).

#### 2.2.3 EEG data pre-processing

EEG data were preprocessed with the EEGLAB toolbox version v2021.1 (Delorme & Makeig, 2004) for MATLAB (version 2022a; MathWorks, Natick, MA, USA). First, to suppress high frequencies, data were low-pass filtered (35 Hz, FIR, hamming window, filter order: 166). Data were downsampled to 250 Hz to reduce computational demands and disk space consumed, and high-pass filtered at (1 Hz, FIR, hamming window, filter order: 826) to suppress very slow trends in the data. Edge artefacts were removed by discarding the first and last 5 s of each recording. Identification of improbable channels was conducted using the EEGLAB extension trimOutlier (https://sccn.ucsd.edu/wiki/EEGLAB_Extensions) with an upper and lower boundary of two standard deviations of the mean standard deviation across all channels (Stroke survivors: *M* = 1.4, *SD* = 0.6, range 0-3 channels; HC *M* = 0.5, *SD* = 0.9, range 0-3 channels). Channels exceeding this threshold were reconstructed using spherical interpolation. At this stage two copies of the dataset were created. One copy was cleaned using Artifact Subspace Reconstruction (ASR, as implemented in EEGLAB plugin clean_rawdata(), (Blum et al., 2019; T. Mullen et al., 2013; T. R. Mullen et al., 2015)) aiming to remove non-stereotypical artefacts to improve Independent Components Analysis (ICA). We used temporal ICA across channels using the runica algorithm (Makeig et al., 1995) to estimate 30 independent components. Components reflecting line noise, eye, muscle and heart activity were identified using the EEGLAB extension ICLabel (Pion-Tonachini et al., 2019) and confirmed by visual inspection. Components flagged as artefactual (Stroke survivors: *M*=6.1, *SD*=2.1, range 4-11 components; HC *M*=5.9, *SD*=2.3, range 4-11 components) were regressed out of the second copy created after spherical interpolation. From here, data were processed differently for task data and resting state data.

For the resting state data, the power spectral density (PSD) estimate was computed with a 2 second window and a 1 second overlap. The spectra were normalised to represent relative power by dividing each bin in the spectrum by the total power across all bins. For each participant, a 2-dimensional array (frequencies × sensors) containing power was obtained.

Task data were segmented from -1.5 to 3.5 s relative to the button press. Artefactual epochs as indicated by the joint probability (EEGLAB function pop_jointprob.m, *SD* = 3) were discarded from further analyses (Stroke survivors: *M* = 11.3%, *SD* = 3.7%, range 5.3-22.1%; HC *M* = 10.3%, *SD* = 3.4%, range 5.9-24.8%). Time-frequency representations of individual trials were calculated using the Fieldtrip functions (www.fieldtriptoolbox.org; (Oostenveld et al., 2010)) integrated in SPM12 using Morlet wavelet analysis with a wavelet width of 9. Power values were baseline corrected (-1 to -0.2 s). Data were averaged across the β-frequency range (13-30 Hz), resulting in two-dimensional array (time x sensors). Stroke survivor data were grouped as affected and unaffected UL, while HC data were grouped as non-dominant and dominant UL. Data from patients with left-hemisphere lesions were mirrored, aligning the right hemisphere with the affected hemisphere and the left hemisphere with the unaffected hemisphere, and the left and right UL with the affected and unaffected UL, respectively.

### 2.3 Statistical analysis

Relationships between clinical outcome measures was assessed using Spearman’s rank correlation, and confidence intervals were computed using permutation bootstrapping with 5,000 iterations. A Bonferroni correction was applied for the two comparisons, resulting in a significance threshold of *p* < 0.025.

Brain activity data were analysed using a General Linear Model (GLM) framework as implemented in glmtools (https://pypi.org/project /glmtools/; (Quinn et al., 2024)). Two GLMs (task, rest) were constructed to compare group averages, each with three regressors: two categorical predictors using dummy coding for participant group (HC/stroke), and two continuous covariate representing z-transformed participant age and time on program (i.e., time between start of the program and the EEG recording). Parameters were estimated using the Moore-Penrose Pseudo-Inverse for each frequency–sensor pair in the resting-state data, resulting in a spectrum of parameters. For the task data, parameters were estimated for each time–sensor pair, yielding a time course of parameters. A contrast of parameter estimates was defined to test for differences between HC and chronic stroke. The resulting contrast estimates (COPEs) and their associated standard errors are used to compute a t-test for this contrast.

In the patient group, parametric models were constructed to assess associations with initial clinical scores and changes on the program. Two models were created for each condition (task and rest), one for clinical scores at admission and one for percentage change. Each model included a constant regressor and two parametric regressors: one for z-transformed clinical scores (FME-UL, CAHAI) or percentage change, and one for time on the programme. Parameters were estimated using the Moore-Penrose Pseudo-Inverse, and contrasts of parameter estimates tested differences from zero. The resulting contrast estimates (COPEs) and standard errors were used for a t-test.

Null-hypothesis testing was conducted using cluster based non-parametric permutation tests (Maris & Oostenveld, 2007; Nichols & Holmes, 2001). This approach makes minimal assumptions, addresses the issue of multiple comparisons, and is suitable when parametric assumptions are not met (Nichols & Holmes, 2001). Row-shuffle permutations were used to compare subgroup differences and test relationships between variables, while sign-flipping permutations were used to test whether a measurement deviated from zero. A null distribution of cluster statistics was generated by permuting the data and calculating the sum of *t*-values within the largest contiguous cluster exceeding a cluster-forming threshold of *t* = 3.5. Observed cluster statistics were compared to this null distribution and considered significant if they exceeded a predefined critical value. In this study, the null distribution was constructed using 5,000 permutations, with the 95th percentile used as the significance threshold. Multiple comparisons were controlled for within each statistical map using a cluster-based permutation approach. No formal correction was applied across the different models.

Outliers in clinical scores and normalised clinical change scores were identified using Grubbs’ test; analyses were repeated with these participants excluded to assess the robustness of the results.

We conducted post hoc sensitivity analyses to quantify the effect sizes detectable with the available sample sizes. For passive movement of the affected UL (N = 39), the study had 80% power to detect correlations of approximately |ρ| ≥ 0.43 at α = 0.05. For passive movement of the unaffected Ul and resting state (N = 31), detectable effects were approximately |ρ| ≥ 0.49. Thus, the absence of significant associations argues against moderate-to-large effects, although smaller effects may remain undetectable.

### 2.4 Data availability

All data produced in the present study are available upon reasonable request to the authors.

## 3. Results

### 3.1 Participant characteristics

A total of 40 individuals with chronic stroke were recruited from the QSUL programme. One dataset was excluded due to a medical event that led the participant to withdraw from the programme, resulting in a final sample of 39 participants. The sample included 31 males and 8 females, with a mean age of 55.6 years (*SD* = 11.6; range 37.2-82.1 years). The left UL was affected in 12 participants, and the right UL was affected in 27 participants. At the time of testing, the mean time since stroke was 2.3 years (*SD* = 1.5; range 0.65-8.25 years). EEG data were obtained on average 5.8 days after starting the QSUL programme (Median = 4.5; *SD* = 4.6; range 0-13 days). Due to time constraints related to clinical workflow, complete datasets were not available for all participants, resulting in 39 datasets for the task data of the affected UL condition, 31 for task data of the unaffected UL, and 31 for resting state data. EEG data were also collected from 26 HC including 9 females and 17 males, with a mean age of 56.5 years (*SD* = 11.1; range 37.7-85.2 years).

### 3.2 Clinically meaningful gains in the upper limb following the QSUL programme

Scores for FM-UE and CAHAI for the affected UL at admission (T1) and discharge (T2) are presented in **Fig. 1** and **Table 1**. Initial FM-UE scores were correlated with initial CAHAI scores (rho(37) = 0.74, 95% bootstrap CI [-0.51, 0.87], *p* < 0.001), and improvements in CAHAI were associated with improvements in FM-UE (rho(37) = 0.59, 95% bootstrap CI [0.35, 0.76], *p* < 0.001). Wilcoxon signed-rank tests demonstrated significant improvements across both measures following the QSUL programme (FM-UE (*p* < 0.001, *z* = -5.24) and CAHAI (*p* < 0.001, *z* = -5.45)). Importantly, gains on both the FM-UE and CAHAI exceeded their respective MCID (Barreca et al., 2005; Page et al., 2012), consistent with previously reported outcomes of the QSUL programme (Ward et al., 2019).

**Fig. 1.**
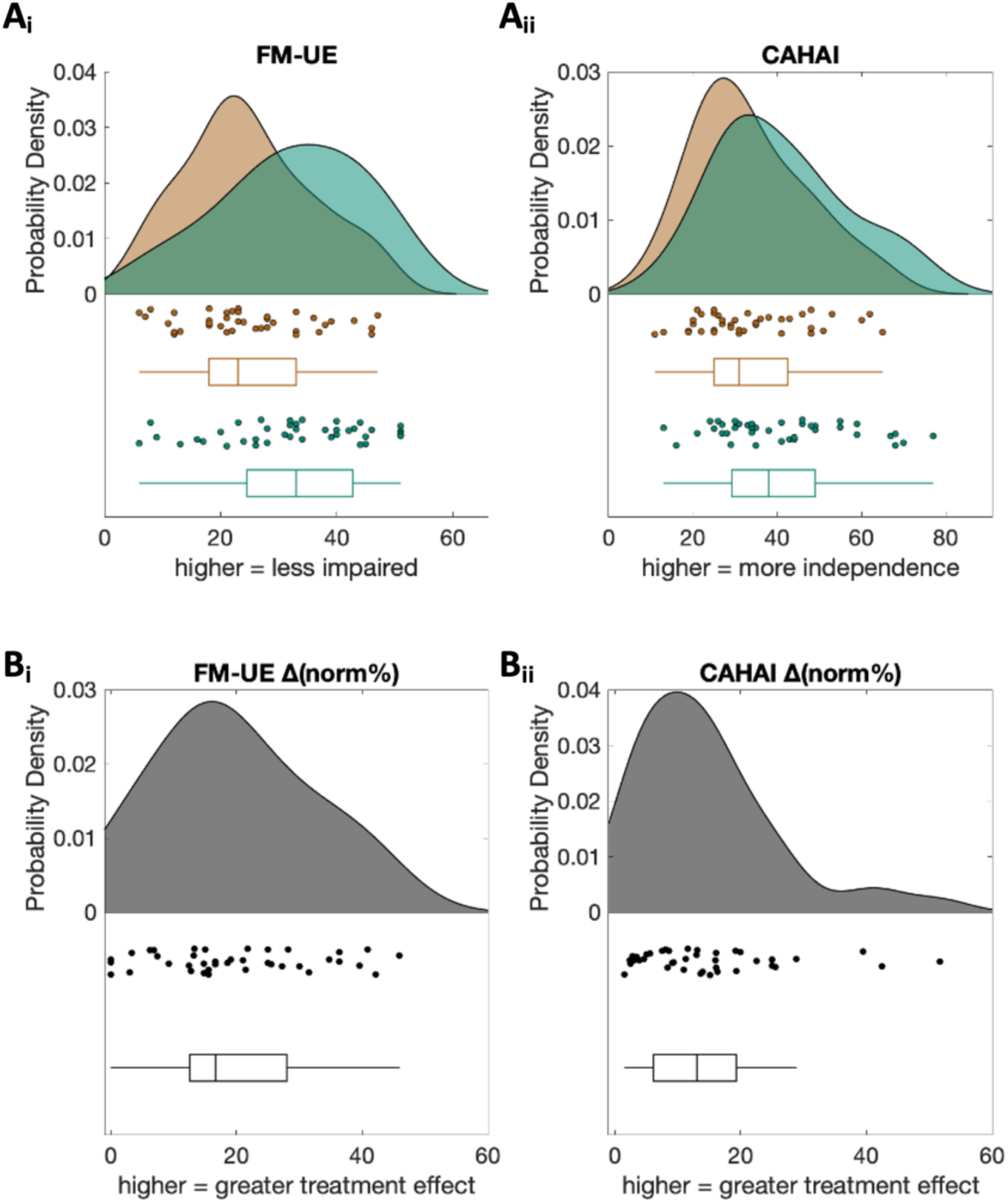
Clinical scores at admission (T1) and discharge (T2) and normalised percent change. **Ai)** FM-UE at admission (brown) and discharge (green). Shown is the probability function, the corresponding individual datapoints and boxplot (median, interquartile range [25th–75th percentiles], whiskers extending to ±1.5× IQR). **Aii)** FM-UE normalised percent change. Shown is the probability function, the corresponding individual datapoints and boxplot (median, interquartile range [25th–75th percentiles], whiskers extending to ±1.5× IQR).. **Bi)** Same as **Ai)**, for CAHAI. **Bii)** Same as **Aii)**, for CAHAI.

**Table 1.**
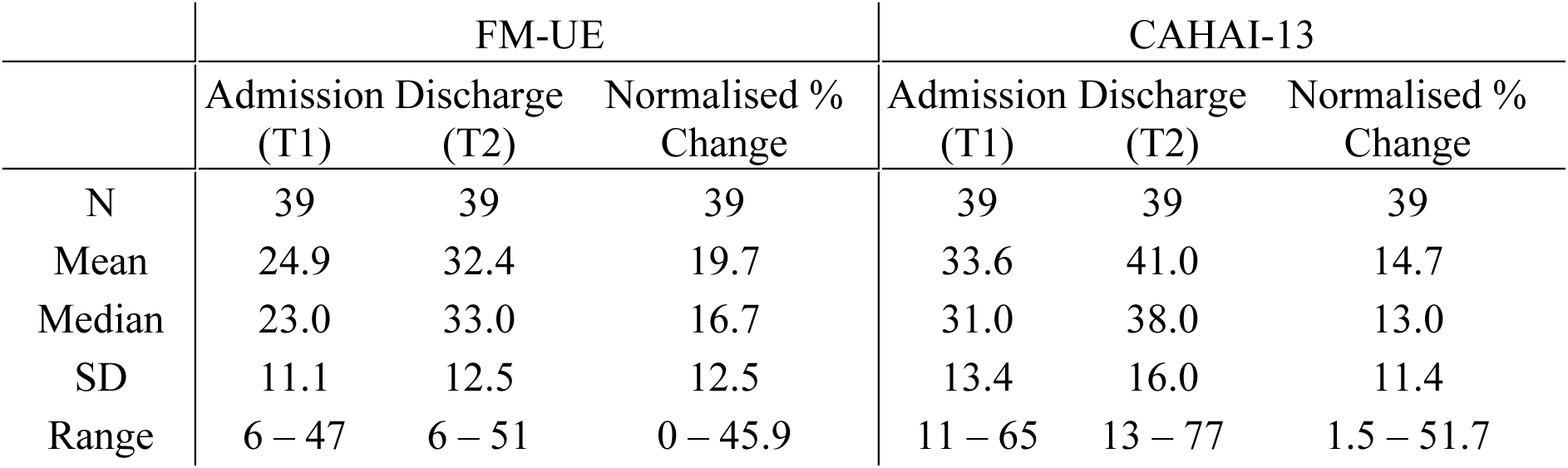
Clinical scores at admission (T1) and discharge (T2) and their normalised percent change.

|  | FM-UE |  |  | CAHAI-13 |  |  |
| --- | --- | --- | --- | --- | --- | --- |
|  | Admission (T1) | Discharge (T2) | Normalised % Change | Admission (T1) | Discharge (T2) | Normalised % Change |
| N | 39 | 39 | 39 | 39 | 39 | 39 |
| Mean | 24.9 | 32.4 | 19.7 | 33.6 | 41.0 | 14.7 |
| Median | 23.0 | 33.0 | 16.7 | 31.0 | 38.0 | 13.0 |
| SD | 11.1 | 12.5 | 12.5 | 13.4 | 16.0 | 11.4 |
| Range | 6 – 47 | 6 – 51 | 0 – 45.9 | 11 – 65 | 13 – 77 | 1.5 – 51.7 |

### 3.3 Stroke-related changes in β-activity

Next, we investigated stroke-related differences in β-activity during passive movement and neural power at rest. In response to passive movement, a clear β-event-related desynchronization/synchronization (β-ERD/ERS) complex was observed in both UL of HC and chronic stroke survivors (**Fig. 2A**, **Fig. 2B**). However, compared to HC, chronic stroke survivors exhibited a reduced β-ERD (less negative) and β-ERS (less positive) responses in both UL.

**Fig 2.**
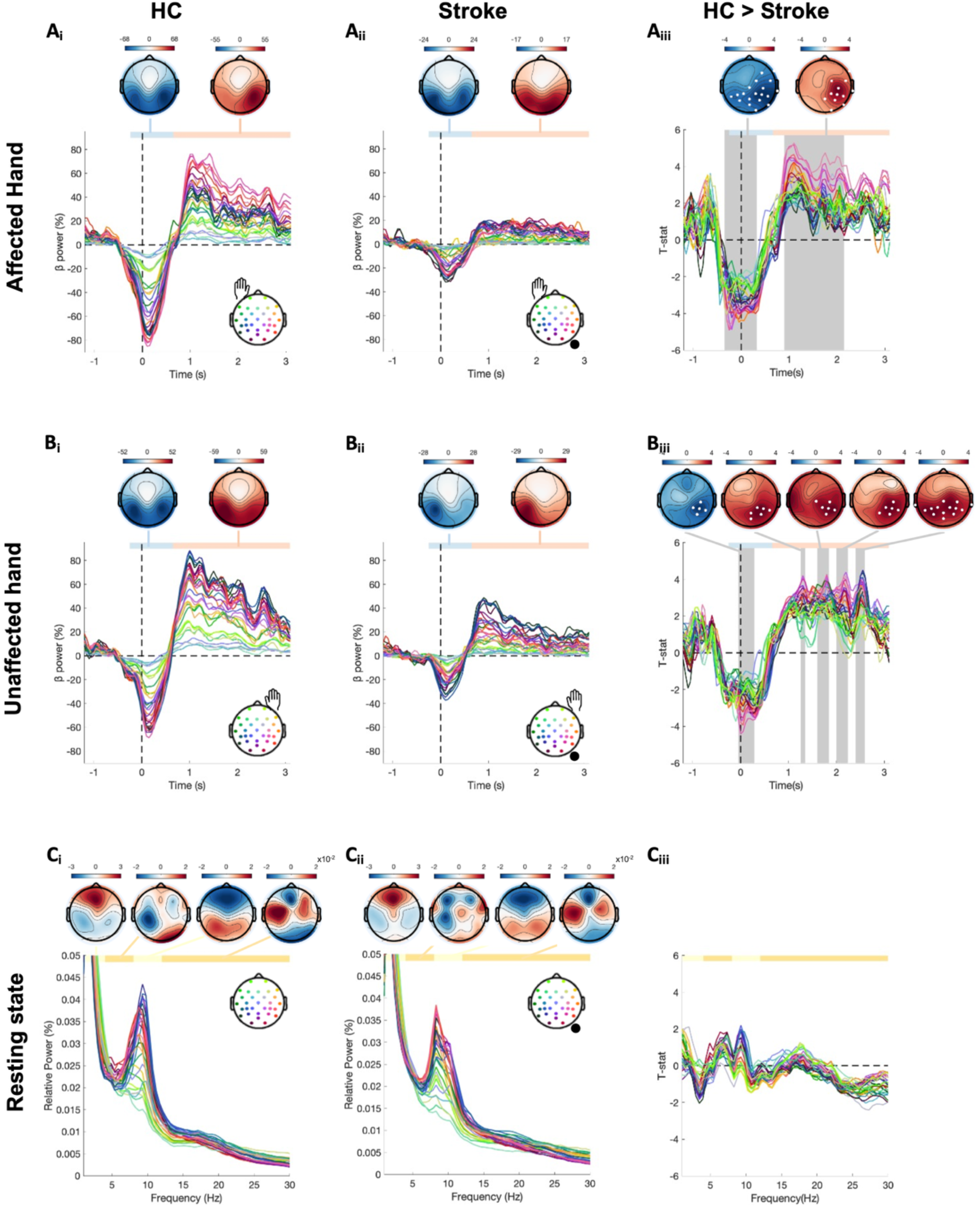
Brain activity in chronic stroke relative to healthy controls. **Ai)** β-power during passive movement for healthy controls (HC) of the non-dominant UL (N = 26). Mean power estimates as a function of time and space. The inlay illustrates the channel location colour coding. ERD and ERS time windows are highlighted in blue and red accordingly. Topographies show the spatial pattern of β-power averaged over ERD and ERS time windows. **Aii)** Same as ai) for passive movement of the nondominant UL in HC vs affected UL in stroke survivors (N = 39). The inlay illustrates the channel location colour coding, the lesioned hemisphere (black circle) and the UL that was moved (here affected UL). **Aiii)** t-values as a function of time and space for the contrast between HC and chronic stroke survivors during passive movement of the affected UL. Statistical significance is assessed by time x channel cluster permutation testing. Significant times are indicated in by the grey area. Significant channels are highlighted in white in the topography. Topographies are scaled from plus and minus the maximum absolute value of the data plotted. **Bi)** Same as Ai) for the dominant UL in HC (N = 26). **Bii)** Same as Aii) for the unaffected UL in stroke survivors (N = 31). **Biii)** Same as Aiii) for dominant UL in HC vs unaffected UL in chronic stroke. **Ci)** Resting state power for HC (N = 26). Mean power estimates as a function of frequency and space. The inlay illustrates the channel location colour coding. δ-, θ-, α-, β-frequency bands are highlighted in different shades of yellow. Topographies on the top show the spatial pattern of power averaged over each frequency band. **Cii)** Same as ci) for stroke survivors (N = 31). The inlay illustrates the channel location colour coding, the lesioned hemisphere (black circle). **Ciii)** t-values as a function of frequency and space for the contrast between HC and chronic stroke survivors. Statistical significance is assessed by frequency x channel cluster permutation testing. No significant difference was found.

Specifically, compared to HC passive movement of the affected UL in chronic stroke showed a significantly attenuated β-ERD (less negative) at -0.35 to 0.35s in a large spatial cluster comprising 19 channels across the contralateral (ipsilesional) but also parts of the ipsilateral (contralesional) parietal and occipital regions (**Fig. 2A_iii_**, *threshold_cluster_forming_* = 3.5, *p_cluster_threshold_* = 0.01, *rank_cluster_* = 1, *T_cluster___sum_* = -534.53, *T_cluster_max_* = 4.91, *p_cluster_* < 0.001, β-ERD difference range = −59.38 to -1.75, mean difference = -35.59). Additionally, compared to HC passive movement of the affected UL in chronic stroke showed a significantly attenuated β-ERS (less positive) at 0.9 to 2.2s (**Fig. 2A_iii_**, *threshold_cluster_forming_* = 3.5, *p_cluster_threshold_* = 0.01, *rank_cluster_* = 2, *T_cluster___sum_* = 450.05, *T_cluster_max_* = 5.28, *p_cluster_* < 0.001, β-ERS difference range = 2.40 to 66.91, mean difference = 43.26). The cluster comprised 12 channels located contralaterally (ipsilesional), highlighting a clear laterality effect toward the lesioned hemisphere - which is also the dominant (i.e., contralateral to the UL) hemisphere during passive movement of the affected UL.

During passive movement of the unaffected UL a similar pattern emerged. Compared to HC, chronic stroke patients exhibited a significantly attenuated β-ERD (less negative) at -0.05 to 0.3s (**Fig. 2B_iii_**, *threshold_cluster_forming_* = 3.5, *p_cluster_threshold_* = 0.01, *rank_cluster_* = 3, *T_cluster___sum_* = - 63.68, *T_cluster_max_* = 4.42, *p_cluster_* < 0.001, β-ERD difference range = -55.14 to -15.34, mean difference = -38.93). The cluster comprised 5 channels located over the ipsilateral (ipsilesional) sensorimotor region, again indicating a pronounced laterality effect toward the lesioned hemisphere - which, in this case, represents the nondominant (i.e., ipsilateral to the UL) hemisphere for passive movement of the unaffected UL. Furthermore, compared to HC passive movement of the unaffected UL in chronic stroke resulted in significantly attenuated β-ERS (less positive) in four clusters (**Fig. 2B_iii_**, *threshold_cluster_forming_* = 3.5, *p_cluster_threshold_* = 0.01; *<u>rank_cluster_</u>* <u>= 1</u>, *T_cluster___sum_* = 118.74, *T_cluster_max_* = 4.50, *p_cluster_* < 0.001, time range = 2.4 to 2.6s, β-ERS difference range = 5.44 to 47.43, mean difference = 33.86; *<u>rank_cluster_</u>* <u>= 2</u>, *T_cluster___sum_* = 83.58, *T_cluster_max_* = 4.39, *p_cluster_* = 0.001, time range = 2 to 2.25s, β-ERS difference range = 22.39 to 45.91, mean difference = 33.13; *<u>rank_cluster_</u>* <u>= 4</u>, *T_cluster___sum_* = 57.08, *T_cluster_max_* = 4.27, *p_cluster_* = 0.002, time range = 1.7 to 1.85s, β-ERS difference range = 8.40 to 48.57, mean difference = 31.64; *<u>rank_cluster_</u>* <u>= 5</u>, *T_cluster___sum_* = 49.55, *T_cluster_max_* = 4.21, *p_cluster_* = 0.002, time range = 1.25 to 1.35s, β-ERS difference range = 34.62 to 58.13, mean difference = 48.09). All clusters were located over the ipsilesional sensorimotor area, except for one cluster that showed a bilateral distribution.

At rest, no significant differences in resting-state power were observed between HC and stroke survivors across any frequency band or recording channel (**Fig. 2C***, threshold_cluster_forming_* = 3.5, *p_cluster_threshold_* = 0.01).

### 3.4 Associations between baseline clinical measures and β-activity

We next investigated whether clinical measures at admission were associated with β-activity during passive movement or neural power at rest. A significant relationship was found between CAHAI scores at admission (T1) and β-activity during passive movement of the unaffected UL within the β-ERD time window, with 16 sensors over contralateral (contralesional) and ipsilateral (ipsilesional) sensorimotor areas, such that higher CAHAI scores (greater activity) were associated with stronger β-ERD (more negative) (*threshold_cluster_forming_* = 3.5, *p_cluster_threshold_* = 0.01; *rank_cluster_* = 1, *T_cluster___sum_* = -203.07, *T_cluster_max_* = 5.17, *p_cluster_* = 0.042, time range = -0.3 to 2.5s, b range = -16.74 to 0.56, b mean = -6.09). No significant relationships were observed between FM-UE scores at admission and β-activity during passive movement of either the affected or unaffected UL, nor with neural power at rest. Additionally, the timing of EEG administration did not emerge as a significant covariate.

### 3.5 Associations between clinical improvement and β-activity

Finally, we examined whether clinical improvements following the QSUL programme, expressed as the normalised percentage change in clinical scores between admission (T1) and discharge (T2), were associated with β-activity during passive movement or neural power at rest. Note that the timing of EEG administration did not emerge as a significant covariate in any of the following analysis. Significant associations were observed for the CAHAI, but not for the FM-UE (*threshold_cluster_forming_* = 3.5, *p_cluster_threshold_* = 0.01). Specifically, greater clinical improvement on the CAHAI was associated with a stronger β-ERS (more positive) response between 0.5 and 2.2s during passive movement of the affected UL. This effect spanned across all 30 channels indicating a global effect (**Fig. 3A_i_**, *threshold_cluster_forming_* = 3, *p_cluster_threshold_* = 0.05, *rank_cluster_* = 1, *T_cluster___sum_* = -1411.50, *T_cluster_max_* = 4.69, *p_cluster_* = 0.009, b range = -1.60 to 26.40, b mean = 10.56).

**Fig 3.**
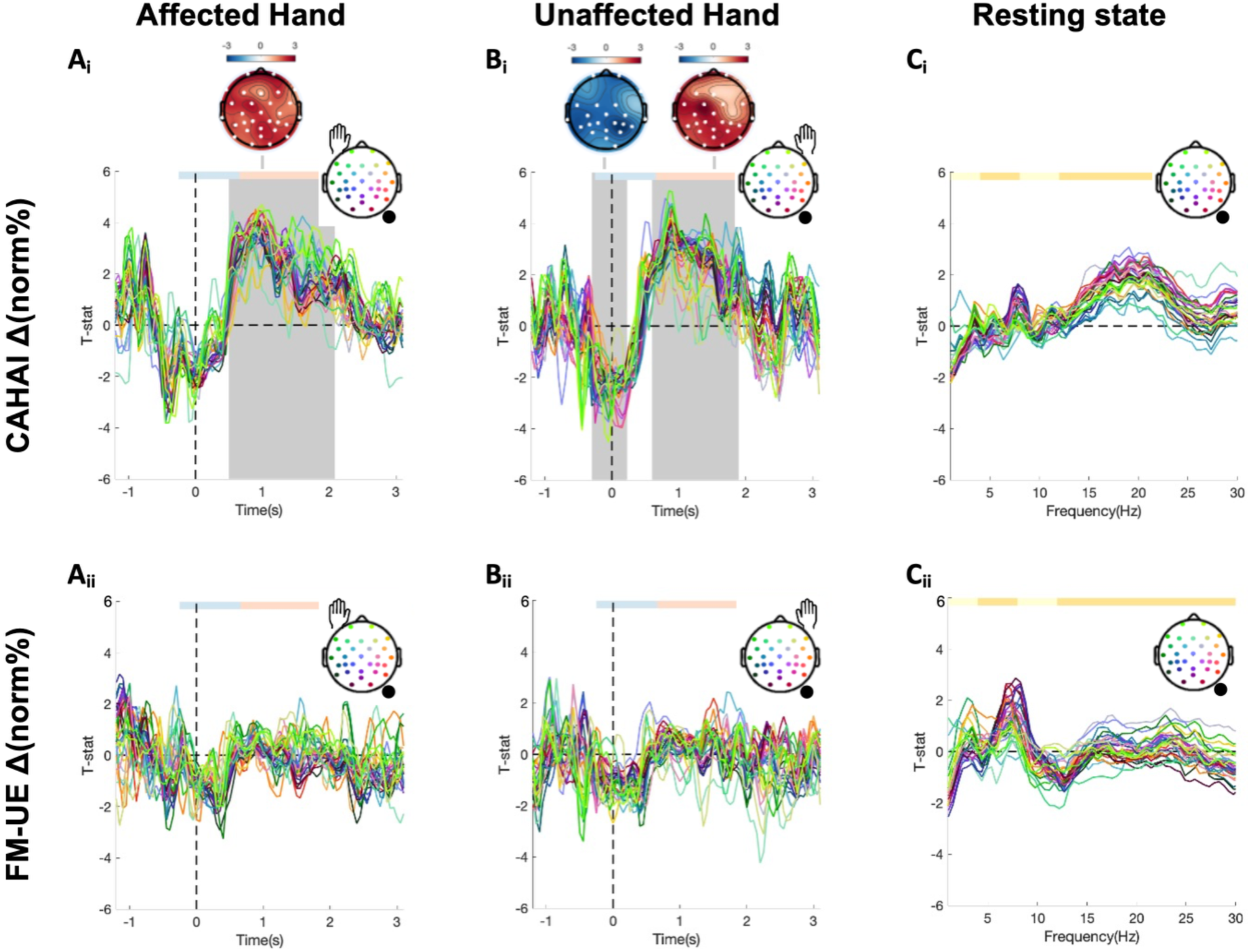
Relationship between β-power during task and rest and changes in motor outcome measures. **A)** t-values as a function of time and space for the contrast correlating β-power during passive movement of the affected UL (N = 39) and CAHAI (top), FM-UE (bottom) change between admission (T1) and discharge (T2). The inlay illustrates the channel location colour coding, the lesioned hemisphere (black circle) and the UL that was moved (here affected UL). ERD and ERS time windows are highlighted in blue and red accordingly. Statistical significance is assessed by time x channel cluster permutation testing. Significant times are indicated by the grey area. Significant channels are highlighted in white in the topography. **B)** Same as a) for β-power during movement of the unaffected UL (N = 31). **C)** t-values as a function of frequency and space for the contrast correlating resting state (N = 31) power and CAHAI (top), FM-UE (bottom) change between admission (T1) and discharge (T2). The inlay topography illustrates the channel location colour coding, the lesioned hemisphere (black circle). δ-, θ-, α-, β-frequency bands are highlighted in yellow. Statistical significance is assessed by frequency x channel cluster permutation testing. No significant associations were found.

During passive movement of the unaffected UL, two temporally distinct associations with CAHAI change were observed. First, greater CAHAI improvement was associated with stronger β-ERD (more negative) between -0.30 and 0.25s over 23 channels (**Fig. 3B_i_**, *threshold_cluster_forming_* = 3, *p_cluster_threshold_* = 0.05, *rank_cluster_* = 2, *T_cluster___sum_* = -266.60, *T_cluster_max_* = 4.51, *p_cluster_* = 0.042, b range = -16.30 to 3.35, b mean = -6.00). Second, greater CAHAI improvement was associated with stronger β-ERS (more positive) between 0.60 and 1.95s over 28 channels (**Fig. 3B_i_**, *threshold_cluster_forming_* = 3, *p_cluster_threshold_* = 0.05, *rank_cluster_* = 1, *T_cluster___sum_* = 1161.40, *T_cluster_max_* = 5.27, *p_cluster_* = 0.011). Notably, the observed associations remained significant after excluding the three patients with large CAHAI changes (**Fig. 1B_ii_**) identified as outliers using Grubbs’ test, indicating that the effects were not driven by extreme values. No significant associations were found between resting-state brain activity and changes in either FM-UE or CAHAI scores (**Fig. 3C***, threshold_cluster_forming_* = 3.5, *p_cluster_threshold_* = 0.01).

## 4. Discussion

In this study, we found that compared to HC, chronic stroke survivors exhibit reduced β-ERD (less negative) and β-ERS (less positive) responses in both UL, highlighting widespread stroke-related alterations in movement-related β-activity. β-power was significantly related to baseline and change scores of the CAHAI, but it was not associated with baseline or change scores of the FMA-UL. These findings suggest that β-activity may serve as a neural correlate of improvement in activity rather than impairment in chronic stroke following intensive UL rehabilitation.

### Movement-related β-activity alterations in the affected and unaffected upper limb

Chronic stroke is associated with alterations in movement-related β-activity. During passive movement of the affected UL, we find reduced β-ERD (less negative) and β-ERS (less positive) in the chronic stroke group compared with HC, indicating attenuated modulation of movement-related β-activity in the ipsilesional hemisphere. This pattern aligns with prior EEG and MEG studies demonstrating diminished movement-related β-activity in the chronic stage post-stroke, and our findings extend this work by providing spatially resolved maps of these effects using a GLM-based framework. For instance, an MEG study by Rossiter et al. (Rossiter et al., 2014) using a hand grip task reported reduced β-ERD (less negative) in chronic stroke patients relative to HC. EEG studies also showed reduced β-ERD (less negative) during various UL motor tasks in chronic stroke survivors (Chalard et al., 2020; Delcamp et al., 2024; Park et al., 2016), reinforcing the robustness of reduced β-activity across cohorts, modalities, and task demands. Kulasingham et al. (Kulasingham et al., 2022) further demonstrated reduced β-ERS (less positive) even in mildly affected chronic stroke survivors. Importantly, similar alterations in movement-related β-activity have also been reported in the acute stage of stroke (Laaksonen et al., 2012; Parkkonen et al., 2018; Zich, Mardell, et al., 2025). While Laaksonen et al. (Laaksonen et al., 2012) and Parkkonen et al. (Parkkonen et al., 2018) observed movement-related β-activity induced by the affected UL beginning to normalize within the first month post-stroke, a reduction relative to HC remains. Taken together with our current findings, this suggests that these alterations emerge early after stroke and persist, at least in part, into the chronic phase.

In this study, we also examined β-activity during movements of the unaffected UL. The unaffected UL is rarely studied in stroke neuroimaging studies, which usually focus exclusively on the affected limb. Exceptions are typically limited to bilateral movement paradigms or mirror therapy designs (e.g., (Rossiter et al., 2015; Tai et al., 2020)). By including the unaffected UL, we were able to assess the limb-specificity of stroke-related changes in movement-related β-activity. Notably, we observed alterations in β-activity during movements of both the affected and unaffected UL, with effects predominantly ipsilesional, indicating that these changes are lateralised and not dependent on which UL is used. Importantly, this pattern of altered β-activity during movements of the unaffected UL has also been reported in acute stroke (Laaksonen et al., 2012; Parkkonen et al., 2018; Zich, Mardell, et al., 2025). While Laaksonen et al. (Laaksonen et al., 2012) and Parkkonen et al. (Parkkonen et al., 2018) observed a normalization of movement-related β-activity induced by the unaffected UL relative to HC, we still observed significant differences between chronic stroke patients and HC in our cohort. These discrepancies may reflect differences in sample characteristics, as our participants likely had more severe strokes, although no formal comparison can be made. In the present study, we did not include a direct measure of motor ability for the unaffected UL, so we cannot relate β-activity to motor performance in the unaffected UL. Nevertheless, evidence from chronic stroke studies indicates that the unaffected UL is not entirely spared. For example, Zhang et al. (L. Zhang et al., 2014) reported that chronic stroke survivors show reduced finger-tapping speed and lower scores in the unaffected UL compared with HC, and Barry et al. (Barry et al., 2020) found persistent deficits in fine motor dexterity, despite preserved grip strength, in the unaffected UL. The reductions in β-ERD (less negative) and β-ERS (less positive) observed in the unaffected UL in our study could therefore reflect these subtle behavioural deficits, although this relationship remains to be tested. Together, these findings suggest that stroke-induced changes extend beyond the affected UL, likely reflecting altered interhemispheric interactions and reorganization of sensorimotor networks. This further supports that even unilateral stroke induces changes across both limbs.

In addition to the motor task data, we also collected resting-state data to evaluate the state-dependency of the observed effects. This allowed us to determine whether stroke-related changes in movement-related β-activity are specific to movement or reflect more general alterations in baseline activity. In the current study, resting-state power did not differ systematically between chronic stroke survivors and HC. Overall, Resting-state activity in chronic stroke is less consistently altered than movement-related β-activity. While some previous studies reported differences in resting-state EEG/MEG activity between chronic stroke survivors and HC - specifically, alterations in θ- and α-power (Snyder et al., 2021; J. J. Zhang et al., 2024) - others found no significant β-power differences compared with HC (Espenhahn et al., 2020). These discrepancies may reflect differences in sample characteristics, such as demographic factors (age, sex) and clinical variables (initial severity, stroke type, lesioned hemisphere), a common issue in stroke research due to typically small sample sizes, or differences in analysis methods. Many previous studies focused on specific frequency bands and electrode sites, relying on a priori assumptions and reducing the data. In contrast, we used a GLM-based, data-driven approach that systematically tests for differences across all frequencies and electrodes, providing an assumption-free evaluation of resting state activity. Thus, as no changes in β-power were observed during rest, the differences in movement-related β-power reported above are unlikely to reflect general alterations in baseline activity.

### β-activity tracks clinical improvements in chronic stroke

Both clinical measures (FM-UE, CAHAI) improved significantly and exceeded their respective minimal clinically important differences over the course of the program, consistent with previous work (Ward et al., 2019). Notably, only changes in CAHAI were significantly associated with movement-related β-activity. Importantly, this association was observed during passive movement of both the affected and unaffected UL, indicating that the β-activity relationship with improvement in activity is not strictly limb-specific but rather reflects a widespread, bilateral cortical effect. This is further supported by the spatial extent of the effect, which spanned numerous channels across both hemispheres, emphasizing the global nature of the neural processes underlying improvement. However, this effect was task-dependent, as no significant relationships were found between β-activity at rest and clinical improvement, suggesting that probing the sensorimotor system is crucial for capturing neural activity linked to UL rehabilitation outcomes.

The CAHAI specifically captures bimanual activity of the UL in real-world tasks, such as opening a jar or pouring water, which require integration of motor planning, coordination, and sensorimotor feedback (Barreca et al., 2005). In contrast, the FM-UE is primarily an assessment of the ability to move out of abnormal UL flexor synergies (Fugl-Meyer et al., 1975). While CAHAI and FM-UE both focus on the UL, the selective correlation between movement-related β-activity and change in CAHAI supports the idea that movement-related β-activity is more closely linked to task performance than to overcoming flexor synergies. This aligns with previous research suggesting that neural activity is a better predictor of improvements in real-world UL use than impairment by itself (Carter et al., 2012; Grefkes & Ward, 2014). Sensorimotor β-activity also plays a crucial role in motor skill acquisition, with studies showing that strengthening of the β-ERD (more negative) and β-ERS (more positive) during practice is linked to motor performance improvements (Barone & Rossiter, 2021; Tan et al., 2014; Wu, Schoenfeld, et al., 2025; Wu, Xu, et al., 2025, 2025; Zich et al., 2018). Thus, alterations in movement-related β-activity may index neural mechanisms that support motor learning in healthy individuals and functional recovery after stroke.

We suggest a tentative model that distinct neural mechanisms may underlie different aspects of post-stroke motor recovery. Movement-related β-activity during passive movement is linked to real-world bimanual UL activity, as measured by the CAHAI, reflecting cortical excitability and inter-regional coordination required for goal-directed actions. Stronger movement-related β-activity is associated with improved activity and responsiveness to rehabilitation, suggesting that modulation of β-activity may reflect the capacity of the sensorimotor system to support motor learning. In contrast, the structural integrity of descending motor pathways such as the corticospinal tract (CST) integrity is more directly associated with motor impairment, as captured by the FM-UE (Byblow et al., 2015; Feng et al., 2015; Schulz et al., 2012; Stinear et al., 2007). Moreover, animal studies have highlighted the importance of spinal cord mechanisms in generating post-stroke UL flexor synergies (Glover et al., 2025), which can be assessed by the FM-UE. Further support comes from recent spinal cord stimulation work showing that projections onto spinal cord motor neurons are needed to reduce motor impairment (Freitas et al., 2025; Powell et al., 2023; Refy et al., 2025). This dual framework of brain function and structure in chronic stroke is further supported by our recent findings in acute stroke survivors (Zich, Mardell, et al., 2025). In that study, structural damage was associated with initial severity but not six-month outcome. In contrast, β-activity was associated with six-month outcome, independent of initial severity. This suggests that similar principles regarding the contribution of β-activity to recovery may apply to both spontaneous recovery in the early stage and rehabilitation-related recovery in the chronic stage. These findings may also indicate that β-activity reflects activity-level engagement, while CST integrity constrains basic motor impairment. As a result, both brain functional activity and structural pathways could be important targets in rehabilitation and serve as biomarkers for recovery outcomes. In doing so, this framework extends previous models that focused primarily on clinical scores and structural information alone (Stinear et al., 2012, 2017).

### Limitations and future directions

We note that the sample size remained relatively small, which may limit the generalisability of the results reported here. Additionally, the number of datasets for the analysis of resting-state data and data from the unaffected UL were lower than the datasets for the affected UL, due to time constraints during data collection. This reflects the inherent challenges of collecting high-quality EEG in a rehabilitation setting, where patient availability and clinical priorities must be carefully balanced. The underrepresentation of female participants in our cohort is a limitation, consistent with trends commonly observed in stroke research (Carcel & Reeves, 2021; Dahlby & Boyd, 2025; Doric et al., 2025; Strong et al., 2021). Further, the timing of EEG data collection varied across participants during the 3-week program, in contrast to the clinical measures, which were consistently collected at admission and discharge of the intervention. Although the timing of EEG administration did not emerge as a significant covariate in the analyses, a more standardized approach would strengthen the design. Ideally, EEG should be collected at fixed time points - at admission to explore its potential as a predictor of therapeutic response, and again at discharge to evaluate changes in neural activity alongside clinical improvements. This approach is being implemented in the related clinical trial (Tedesco Triccas et al., 2025). Although these limitations should be considered, the dataset remains valuable for developing new hypotheses and guiding future research.

## Acknowledgments

Special thanks to all study participants. Thanks to the clinical team, the occupational therapist, physiotherapists, and rehabilitation assistants at The National Hospital for Neurology and Neurosurgery (NHNN), Queen Square, for their dedication in delivering the rehabilitation program and the methodically assessment of the outcome measures.

## Author contributions

CZ: Conceptualization, Methodology, Formal analysis, Investigation, Data Curation, Writing - Original Draft, Visualization, Project administration

LTT: Resources, Writing - Review & Editing

SP: Resources, Writing - Review & Editing

MC: Resources, Writing - Review & Editing

SB: Writing - Review & Editing, Funding acquisition

NSW: Conceptualization, Resources, Writing - Review & Editing, Funding acquisition

